# Ethnic Differences in the Timing and Incidence of Childhood Health Conditions: Evidence from the Born in Bradford Cohort

**DOI:** 10.64898/2026.03.31.26349839

**Authors:** Gillian Santorelli, Rachael W. Cheung, Sunil Bhopal, John Wright

**Author notes:** Correspondence to Gillian Santorelli.

## Abstract

**Objective:** To examine ethnic differences in the incidence and age-related trajectories of childhood health conditions from birth to adolescence within a UK birth cohort.

**Design:** Longitudinal population-based birth cohort with linkage to primary care electronic health records.

**Setting:** Born in Bradford (BiB), a multi-ethnic birth cohort in Bradford, UK.

**Participants:** 13,282 children (36% White British, 44% Pakistani British, 20% other ethnicity) born 2007-2011 with linked primary care records and ≥1 year follow-up.

**Main outcome measures:** Incident diagnoses of atopic conditions (asthma, eczema, allergic rhinoconjunctivitis), overweight/obesity, common mental health disorders (anxiety, depression), and neurodevelopmental disorders (including ADHD and autism). Incidence rates, Kaplan-Meier cumulative incidence, and Cox regression hazards ratios (HRs) were estimated.

**Results:** Atopic conditions emerged early (median onset 5-6 years) and were more common among Pakistani British children, with higher hazards of eczema (HR□2.29, 95%□CI□2.01–2.61), allergic rhinoconjunctivitis (HR 2.27, 2.00-2.58), and asthma (HR□1.35, 1.22–1.50). Overweight/ obesity developed later (median 9-10 years) and were also more frequent in Pakistani British children (HR□1.25, 1.16–1.35). In contrast, common mental health disorders emerged predominantly in early adolescence (median ∼13 years), and both mental health and neurodevelopmental diagnoses were more frequently recorded among White British children; Pakistani British children had lower hazards of neurodevelopmental diagnoses (HR□0.28, 0.23–0.35) and mental health disorders (HR 0.53, 0.41-0.70).

**Conclusions:** Ethnic differences in childhood health are condition-specific and vary by age of onset, emerging at distinct stages. These findings inform the timing of prevention, service planning, and research into underlying mechanism.

**What is already known on this topic:**

- Ethnic differences in childhood health have been previously documented in the UK, including differences in asthma, obesity, and neurodevelopmental and mental health diagnoses.
- South Asian children have been reported to have higher adiposity at lower BMI thresholds and elevated cardiometabolic risk.
- Most existing studies are cross-sectional or focus on single conditions, providing limited insight into when disparities first emerge and how they change across childhood.

**What this study adds:**

- Using longitudinal primary care data from a large UK birth cohort (Born in Bradford), this study characterises the age of onset of multiple major childhood conditions within the same population of White British and Pakistani British children.
- Pakistani British children experience higher incidence of atopic conditions emerging in early childhood (around ages 5-6 years) and overweight and obesity emerging in mid-childhood (around ages 9-10 years).
- In contrast, White British children have higher recorded rates of neurodevelopmental diagnoses across childhood and common mental health disorders emerging predominantly in early adolescence (around age 13 years).
- Ethnic differences in childhood health are condition-specific and vary by age of onset, emerging at distinct stages rather than uniformly across childhood.

**How this study might affect research, practice or policy:**

- Identifies critical developmental windows for intervention: early childhood for atopic disease and mid-childhood for obesity.
- Highlights the need to examine equity in access to neurodevelopmental and mental health assessment, as lower recorded rates may not reflect lower need.
- Provides longitudinal evidence to inform service planning and the timing of prevention strategies across childhood.
- Supports future research to understand the biological, social, and healthcare-related mechanisms underlying ethnic variation in childhood health.

## Introduction

Childhood is a critical window during which biological susceptibility, social context, and environmental exposures shape long-term health trajectories^1,2^. Life-course epidemiology emphasises how early exposures influence later health through sensitive periods and cumulative risk processes^1^. Many chronic conditions first manifest in early life^2^, and often persist into adolescence and adulthood, contributing to substantial long-term disease burden. Understanding when health disparities emerge is therefore important: early divergence may reflect exposures during sensitive periods, whereas widening gaps later may indicate cumulative processes^1^.

In the UK, ethnic differences in child health have been documented^3,4^, including disparities in asthma, adiposity, mental health, and neurodevelopmental diagnoses^5–9^. However, much of this evidence is cross-sectional or condition-specific, limiting understanding of when these disparities emerge and how they evolve across childhood. These differences likely reflect structural, socioeconomic, environmental, and intergenerational influences rather than innate variation^4,10^. Longitudinal birth cohorts linked to routine healthcare data provide a valuable framework for examining these temporal dynamics by identifying when diagnoses occur and how disparities develop over time.

The Born in Bradford (BiB) study is a large, multi-ethnic birth cohort in a socioeconomically deprived and ethnically diverse UK city^11^. Linkage to primary care electronic health records (EHRs) enables longitudinal follow-up across childhood, providing a platform to examine ethnic differences in incident diagnoses within a shared healthcare context.

This study aimed to characterise the timing and magnitude of ethnic differences in childhood disease incidence. We estimated the incidence across major conditions, compared rates between White British and Pakistani British children, examined age-related patterns using cumulative hazard trajectories, and contextualised findings using primary care utilisation.

## Methods

### Study design and population

We used data from the Born in Bradford (BiB) birth cohort, comprising children born in Bradford, UK, between 2007 and 2011^11^. Children were eligible if they had linked primary care records. Of the 13,779 liveborn children, 311 had no primary care linkage and 20 were of unknown ethnicity and were excluded. We further excluded children with less than one year of follow-up (n=166). Written informed consent was obtained from participating mothers, and ethical approval was granted by Bradford National Health Service Research Ethics Committee (ref 06/Q1202/48).

### Data sources

Primary care data were obtained from SystmOne (TPP, Leeds, UK), the sole HER system used by GP practices in the study area. Diagnoses were recorded using the CTV3 (Clinical Terms Version 3) codes. Anthropometric data were integrated from BiB research assessments, routine primary care measurements, and the National Child Measurement Programme (NCMP)^12^. As a contextual indicator of healthcare utilisation, we summarised age-specific rates of primary clinical consultations by ethnicity (consultations per 1,000 person-years, with one consultation per child per date retained to avoid double-counting within-visit coding). To address potential sources of bias inherent in routine healthcare data, we used validated diagnostic code lists and age-based definitions to improve specificity of case ascertainment. As differences in healthcare utilisation may influence recorded diagnoses, we examined age-specific consultation rates by ethnicity to contextualise findings. Analyses were descriptive and adjusted for sex; residual confounding by socioeconomic and environmental factors is possible.

### Outcome Definitions

We examined four thematic groups of health outcomes, defined using prespecified diagnostic code lists (Supplementary Tables 1 - 6).

**1. Atopic conditions**: We grouped asthma with eczema and allergic rhinoconjunctivitis as “atopic conditions” to reflect common clinical and epidemiological clustering. However, we recognise that asthma is a heterogeneous condition and not all cases are atopic in origin. Case definitions followed established age-based criteria to improve specificity^13^.

- Asthma: Presence of ≥1 relevant code recorded at age ≥3 years.
- Eczema: Presence of ≥1 relevant code recorded at age ≥1 year.
- Allergic rhinoconjunctivitis: Presence of ≥1 relevant code at any age.
**2. Adiposity**: Body Mass Index (BMI) was derived from height and weight measurements recorded in BiB, NCMP, or primary care. Age-and sex-adjusted BMI z-scores were calculated using the UK90 growth reference^14^. Analysis was restricted to children aged ≥2 years.

- Overweight (including obesity): First recorded BMI z-score ≥1.04 (≥85th centile)
- Obesity only was defined as a BMI z-score of ≥1.64 (≥95th centile).

For cumulative incidence, children were classified as cases at the time of first recorded threshold crossing, irrespective of subsequent weight trajectory. Because BMI was measured at discrete time points, stepwise increases in cumulative incidence likely reflect measurement opportunities rather than abrupt biological changes in obesity risk.

**3. Common Mental Health Disorders (CMHD)**: Age-appropriate code lists for anxiety and depression by refining existing research lists^15^. Adult-specific codes contexts (e.g. dementia) were removed. Incidence was defined as the first qualifying CTV3 code recorded from age 6 years onwards, to capture meaningful disorders rather than transient development behaviour^16,17^. A small number of implausibly early diagnoses (n=7 before age 6, including three before age 1), supporting this restriction.
**4. Neurodevelopmental disorders**: This composite included attention-deficit/hyperactivity disorder (ADHD); autism spectrum disorder (ASD); intellectual disability; global developmental delay; and specific developmental disorders (speech/language/motor). A case was defined by the presence of any relevant code at any age.

### Exposure

Ethnicity was categorised as White British, Pakistani British, and ‘other’ ethnic groups, based on education or healthcare records. White British and Pakistani British ethnicities represent the majority of the BiB cohort and the Bradford population.

### Follow-up and Censoring

Children were followed from birth until the earliest of first recorded diagnostic code for the outcome of interest; death; deregistration from the GP practice, or the study end date (August 2024). For adiposity outcomes, follow-up was censored at the date of the last available BMI measurements if no overweight/obesity event occurred.

## Statistical Analysis

Baseline characteristics were summarised for the full cohort and stratified by White British and Pakistani British ethnicity. Incidence rates (IRs) per 1,000 person-years were calculated for all conditions. Kaplan-Meier failure functions (1 – S(t)) were used to visualise the cumulative emergence of conditions from birth through adolescence. Cox proportional hazards models estimated Hazard Ratios (HR) comparing Pakistani British children to White British children. As the aim was descriptive rather than causal, models were adjusted for sex. Analyses were conducted in Stata version 17 (StataCorp, Tx) and R version 2024.04.2.

## Results

### Cohort characteristics

A total of 13,282 children met the inclusion criteria, including 4,818 (36.3%) White British, 5,878 (44.3%) Pakistani British, and 2,586 (19.5%) children of other ethnic groups. The mean duration of follow-up was 14.1 years (SD 3.8). Baseline characteristics differed between ethnic groups (Table 1). Pakistani British children were more likely to live in the most deprived IMD quintile, whereas maternal smoking during pregnancy was more common among White British children. Other perinatal factors were broadly comparable.

**Table 1:** Baseline Characteristics.

| **Variable/Category** | **Total Cohort** | **White British** | **Pakistani** |
| --- | --- | --- | --- |
| **Child's sex** |  |  |  |
| Male | 6858 (51.6%) | 2497 (51.8%) | 3006 (51.1%) |
| Female | 6424 (48.4%) | 2321 (48.2%) | 2872 (48.9%) |
| **Ethnicity** |  |  |  |
| White British | 4,818 (36.3%) |  |  |
| Pakistani British | 5,878 (44.3%) |  |  |
| Other ethnicity | 2,586 (19.5%) |  |  |
| **Mode of delivery** |  |  |  |
| Vaginal | 10,020 (76.9%) | 3,562 (75.5%) | 4,519 (77.9%) |
| Caesarean | 3,012 (23.1%) | 1,157 (24.5%) | 1,280 (22.1%) |
| **Type of birth** |  |  |  |
| Singleton | 12937 (97.4%) | 4696 (97.5%) | 5708 (97.1%) |
| Multiple (2/3) | 345 ( 2.6%) | 122 ( 2.5%) | 170 ( 2.9%) |
| **Preterm birth (<37 weeks)** |  |  |  |
| No | 12215 (93.7%) | 4412 (93.5%) | 5467 (94.3%) |
| Yes | 817 ( 6.3%) | 307 ( 6.5%) | 332 ( 5.7%) |
| **Low birthweight (<2500g)** |  |  |  |
| No | 11919 (91.7%) | 4404 (93.7%) | 5233 (90.4%) |
| Yes | 1078 ( 8.3%) | 296 ( 6.3%) | 553 ( 9.6%) |
| **IMD 2010 quintile** |  |  |  |
| 1 | 8990 (67.7%) | 2484 (51.6%) | 4672 (79.5%) |
| 2 | 2363 (17.8%) | 1056 (21.9%) | 845 (14.4%) |
| 3 | 1367 (10.3%) | 817 (17.0%) | 322 ( 5.5%) |
| 4/5 | 556 ( 4.2%) | 459 ( 9.5%) | 37 ( 0.6%) |
| **Smoked during pregnancy** |  |  |  |
| No | 9172 (83.5%) | 2818 (67.5%) | 4728 (96.6%) |
| Yes | 1806 (16.5%) | 1356 (32.5%) | 164 ( 3.4%) |
| **Maternal education** |  |  |  |
| No Degree | 8211 (74.8%) | 3321 (79.6%) | 3617 (73.9%) |
| Degree level | 2760 (25.2%) | 852 (20.4%) | 1275 (26.1%) |
| **Mother's age at** | 27.5 (5.6) | 26.9 (6.1) | 28.0 (5.1) |
| **delivery (Mean, SD)** |  |  |  |
| **Follow-up time**<br>**(years) (Mean, SD)** | 14.2 (3.5) | 13.9 (3.8) | 14.7 (2.9) |

IMD=Index of Multiple Deprivation

### Primary care consultation rates (contextual analysis)

Consultation rates were highest in infancy, declined rapidly in early childhood, before stabilising thereafter (Figure 1). Pakistani British children had modestly higher consultation rates than White British children across most ages. Overall, healthcare utilisation patterns were similar, with no evidence of lower primary care contact among Pakistani British children.

**Figure 1:**
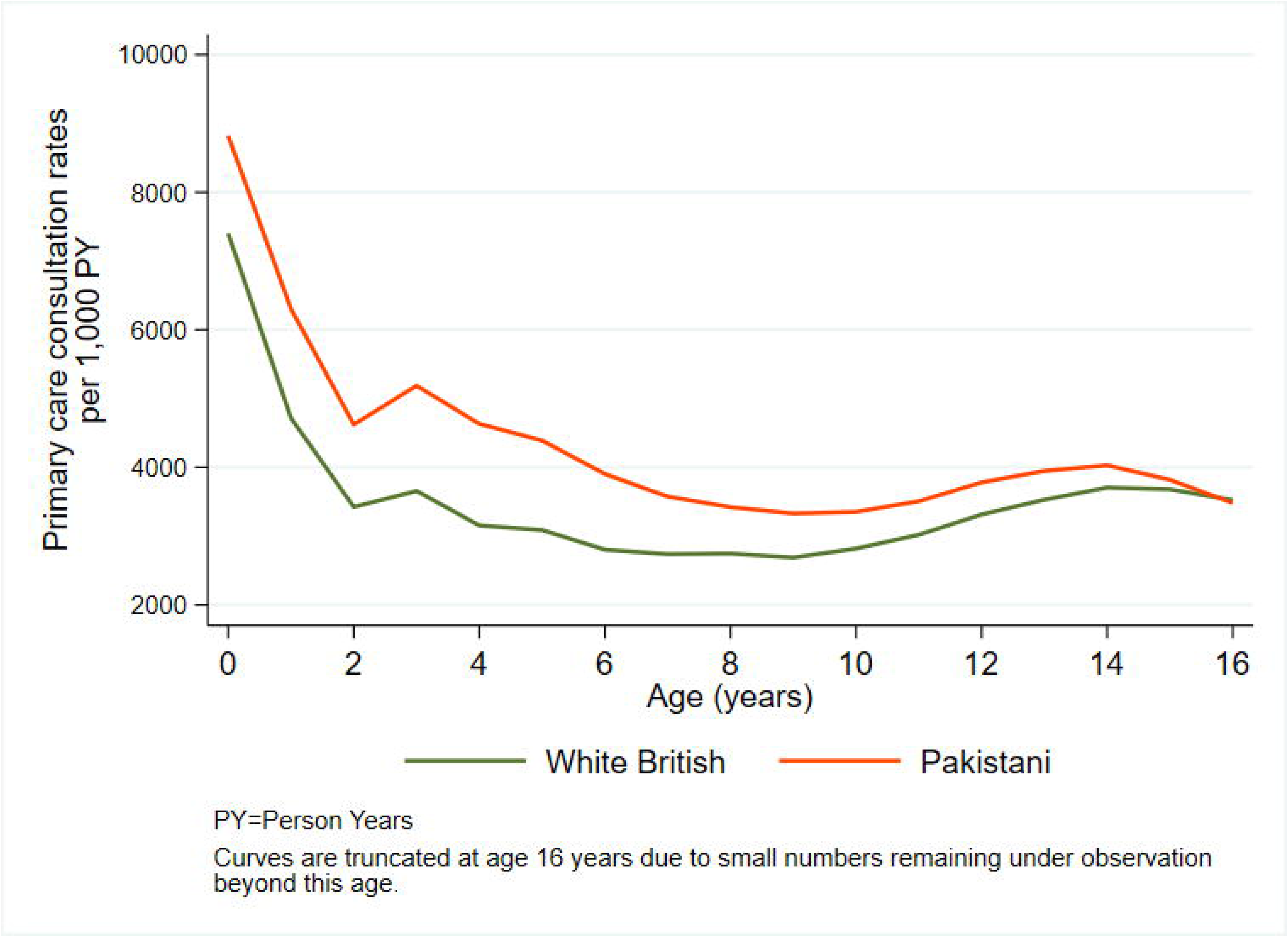
Age-specific primary care consultation rates by ethnicity.

### Incidence and Timing of Conditions

Incidence rates, median age diagnosis, and cumulative risk are summarised in Table 2. Kaplan-Meier curves are shown in Figure 2, hazard ratios in Figure 3, and numbers at risk in Supplementary Table 7.

**Figure 2:**
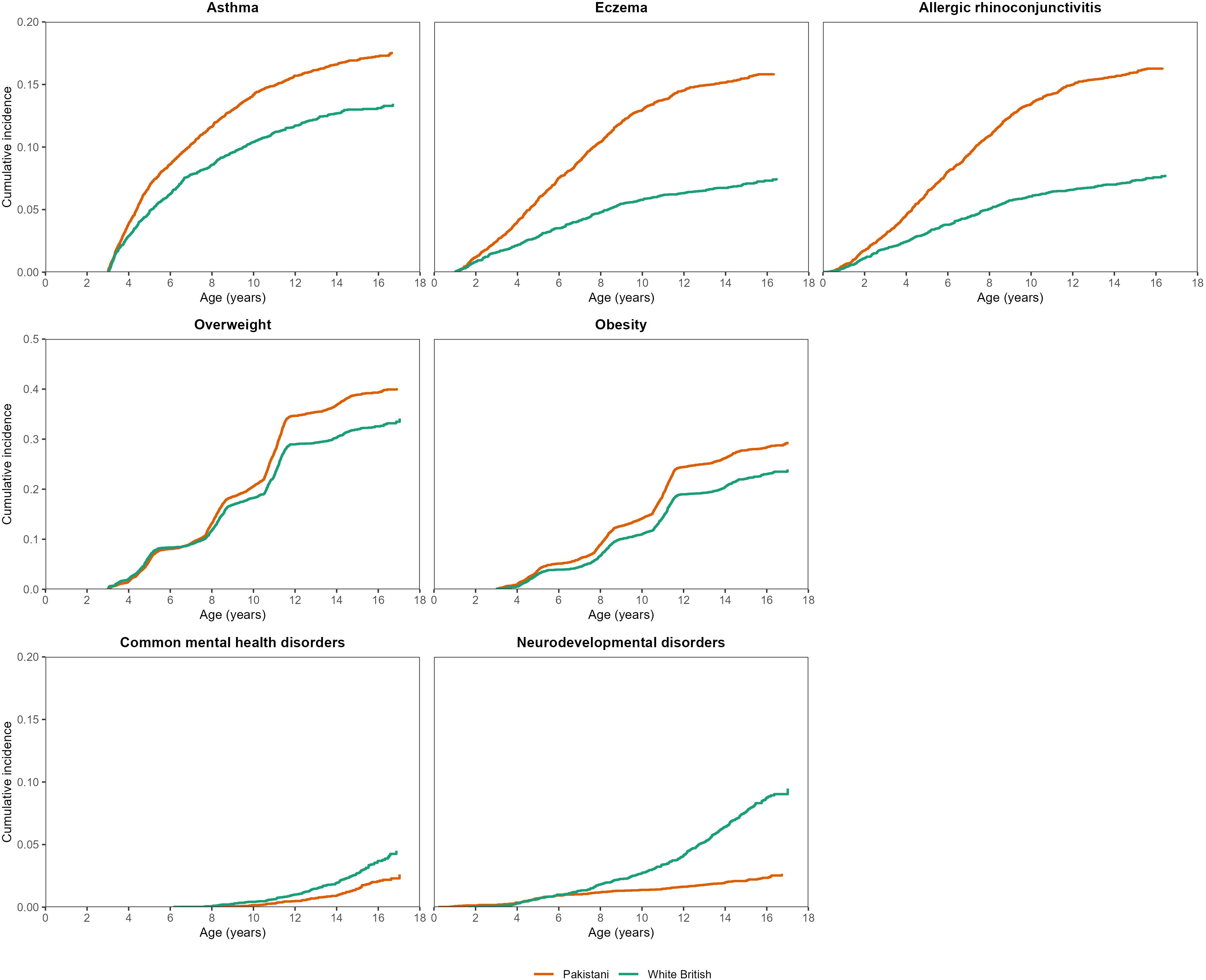
Cumulative incidence of childhood conditions by ethnicity.

**Figure 3:**
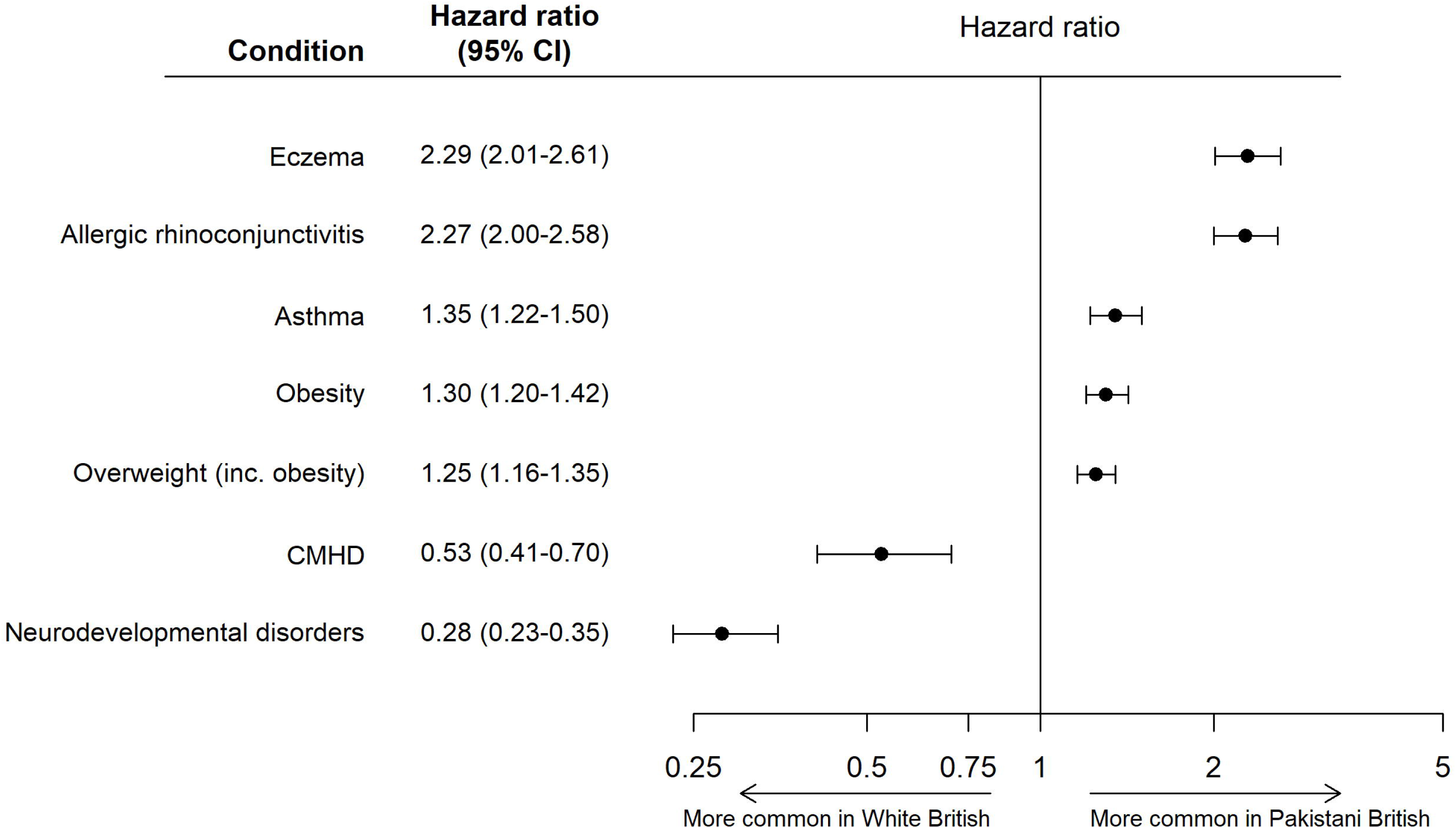
Hazard ratios for childhood conditions by ethnicity. CMHD = Common Mental Health Disorders.

**Table 2.** Incidence, timing of diagnosis, and cumulative risk of childhood health conditions.

| Group | Cases | Person-Years | IR/1,000 Person Years | 95% CI | Median age at diagnosis | Cumulative Incidence (%) |  |  |
| --- | --- | --- | --- | --- | --- | --- | --- | --- |
|  |  |  |  |  |  | Age 5 | Age 10 | Age 15 |
| Asthma (Age 3+) |  |  |  |  |  |  |  |  |
| Overall | 1855 | 133,775.0 | 13.87 | (13.25-14.51) | 5.86 | 5.9 | 12.3 | 14.9 |
| White British | 569 | 47,889.2 | 11.88 | (10.94-12.90) | 6.02 | 4.8 | 10.4 | 13.0 |
| Pakistani | 958 | 60,575.2 | 15.82 | (14.84-16.85) | 5.88 | 6.9 | 14.2 | 16.9 |
| Eczema (Age 1+) |  |  |  |  |  |  |  |  |
| Overall | 1509 | 161,516.5 | 9.34 | (8.88-9.83) | 6.09 | 4.7 | 10.1 | 12.2 |
| White British | 313 | 59,094.6 | 5.30 | (4.74-5.92) | 5.74 | 2.8 | 5.8 | 7.1 |
| Pakistani | 871 | 72,191.8 | 12.07 | (11.29-12.89) | 6.09 | 5.8 | 13.0 | 15.5 |
| Allergic Rhinoconjunctivitis |  |  |  |  |  |  |  |  |
| Overall | 1567 | 174,888.5 | 8.96 | (8.53-9.41) | 5.84 | 5.1 | 10.5 | 12.6 |
| White British | 327 | 63,954.0 | 5.11 | (4.59-5.70) | 5.57 | 3.1 | 6.0 | 7.4 |
| Pakistani | 903 | 78,091.0 | 11.56 | (10.83-12.34) | 5.92 | 6.4 | 13.4 | 16.0 |
| Overweight (inc. obesity) (Age 2+) |  |  |  |  |  |  |  |  |
| Overall | 3315 | 103,533.6 | 32.02 | (30.95-33.13) | 8.85 | 6.1 | 19.5 | 35.7 |
| White British | 1014 | 35,857.9 | 28.28 | (26.59-30.07) | 8.61 | 6.6 | 18.2 | 31.9 |
| Pakistani | 1718 | 48,675.4 | 35.30 | (33.66-37.00) | 9.41 | 5.8 | 20.6 | 38.9 |
| Obesity (Age 2+) |  |  |  |  |  |  |  |  |
| Overall | 2653 | 125,532.3 | 21.13 | (20.34-21.95) | 9.59 | 3.3 | 12.9 | 25.1 |
| White British | 821 | 45,038.8 | 18.23 | (17.02-19.52) | 9.77 | 3.0 | 11.0 | 22.1 |
| Pakistani | 1371 | 57,783.7 | 23.73 | (22.50-25.02) | 9.78 | 3.8 | 14.1 | 27.8 |
| **Common Mental Health Disorders (Age 6+)** |  |  |  |  |  |  |  |  |
| **Overall** | **260** | **110,848.0** | **2.35** | **(2.08-2.65)** | **13.31** | † | **0.2** | **2.0** |
| White British | 126 | 38,682.7 | 3.26 | (2.74-3.88) | 13.09 | † | 0.4 | 2.7 |
| Pakistani | 92 | 51,460.0 | 1.79 | (1.46-2.19) | 13.53 | † | 0.2 | 1.5 |
| **Neurodevelopmental Disorders** |  |  |  |  |  |  |  |  |
| **Overall** | **517** | **186,105.4** | **2.78** | **(2.55-3.03)** | **10.77** | **0.7** | **1.8** | **4.1** |
| White British | 330 | 65,436.1 | 5.04 | (4.53-5.62) | 11.73 | 0.6 | 2.7 | 7.6 |
| Pakistani | 126 | 85,314.1 | 1.48 | (1.24-1.76) | 7.53 | 0.7 | 1.4 | 2.1 |

† Person-years not applicable for common mental health disorders at age 5 due to case definition ≥6 years.

### Atopic Conditions

Atopic conditions were common and typically diagnosed in early childhood, with a median age at diagnosis of approximately 5-6 years. Incidence rates were higher among Pakistani British children than White British children across all atopic outcomes, including asthma (15.8 vs 11.9 per 1,000 person-years (PY)), eczema (12.1 vs 5.3 per 1,000 PY), and allergic rhinoconjunctivitis (11.6 vs 5.1 per 1,000 PY). Pakistani British children also had higher hazards of atopic disease, with more than double the hazard of eczema (HR 2.29; 95% CI: 2.01–2.61) and allergic rhinoconjunctivitis (HR 2.27; 95% CI: 2.00–2.58), and a more modest increase for asthma (HR 1.35; 95% CI: 1.22–1.50). Cumulative incidence curves showed that ethnic differences emerged early and widened through childhood. By age 15 years, cumulative risk was higher among Pakistani British children for asthma (16.9% vs. 13.0%), eczema (15.5% vs. 7.1%), and allergic rhinoconjunctivitis (16.0% vs. 7.4%).

### Adiposity

Adiposity outcomes were also common but arose later, with median ages at onset of approximately 9-10 years. Incidence rates were higher among Pakistani British children than White British children for both overweight (including obesity) (35.3 vs. 28.3 per 1,000 PY) and obesity (24.7 vs 18.2 per 1,000 PY). Pakistani British children had a higher hazard of developing overweight or obesity (HR 1.25; 95% CI: 1.16–1.35). Cumulative incidence increased from mid-childhood in both groups, with persistent differences over time. By age 15 years, cumulative risk of being overweight (including obesity) was 38.9% among Pakistani British children compared with 31.9% among White British children; corresponding risks for obesity was 27.8% and 22.1%.

### Common Mental Health Disorders

Common mental health disorders (CMHD) were relatively uncommon, with an overall incidence rate of 2.4 per 1,000 PY. Incidence rates were higher among White British children (3.3 per 1,000 PY) than among Pakistani British children (1.8 per 1,000 PY). These conditions emerged predominantly in early adolescence, with a median age of diagnosis of approximately 13 years. Pakistani British children had a lower hazard of CMHD diagnosis than White British children (HR 0.53; 95% CI: 0.41-0.70). Cumulative incidence remained low throughout childhood, increasing from around age 10. By 15 years, cumulative risk was 2.7% among White British children and 1.5% among Pakistani British children.

### Neurodevelopmental Conditions

Neurodevelopmental conditions were relatively uncommon overall (2.8 per 1,000 PY) but showed marked ethnic differences. Incidence rates were substantially higher among White British children (5.0 per 1,000 PY) than among Pakistani British children (1.5 per 1,000 PY). The median age at diagnosis was 10.8 years overall, with earlier diagnosis in Pakistani British children (7.5 years) than White British children (11.7 years). Despite this, Pakistani British children had a markedly lower hazard of neurodevelopmental diagnosis (HR 0.28; 95% CI: 0.23–0.35). Cumulative incidence increased gradually over childhood, reaching 7.6% among White British children and 2.1% among Pakistani children by age 15. The composite outcome was predominantly composed of socioemotional and behavioural diagnoses (82%), consistent with autism spectrum disorder and ADHD (Supplementary Table S8).

## Discussion

### Principal Findings

In this longitudinal birth cohort, we observed clear age-patterned ethnic differences across multiple domains of childhood morbidity. Atopic conditions emerged early and were more common among Pakistani British children, with disparities evident from infancy and widening through adolescence. Adiposity outcomes arose later, with higher risk among Pakistani British children from mid-childhood onwards. In contrast, common mental health disorders emerged in early adolescence, and, together with neurodevelopmental diagnoses, were more frequently recorded among White British children. Overall, these findings demonstrate that ethnic differences in childhood health vary by condition and timing, emerging at distinct stages rather than following a single trajectory.

### Comparison With Existing Literature

#### Atopic conditions

Our findings align with UK EHR studies reporting higher incidence of allergic disease in South Asian populations than White groups. National analyses have shown higher incidence of asthma, allergic rhinitis, and eczema among South Asian children^18,19^, and previous Born in Bradford work has demonstrated greater early-life burden of eczema and rhinitis in Pakistani-origin children^20^. Earlier UK studies suggested lower symptom-based asthma prevalence in South Asian children, but these relied mainly on cross-sectional parental reports of wheeze ^21^. Differences in outcome definition, healthcare utilisation, and diagnostic practice may explain the divergence from more recent incidence-based findings. Previous BiB work has also shown that parent-reported and GP-recorded allergic disease capture overlapping but non-identical constructs^22^. Ethnic differences may therefore partly reflect differences in symptom-recognition, help-seeking, or diagnostic recording, in addition to biological susceptibility. At the same time, evidence suggests that genetic susceptibility and gene-environment interactions contribute to variation in atopic disease across populations, including differences in skin barrier function and immune response pathways, may also contribute^23^. Our grouping of asthma with other atopic conditions reflects a pragmatic epidemiological approach; however, we acknowledge that not all asthma is atopic, and this may introduce some heterogeneity into this outcome group.

#### Adiposity

The higher risk of overweight and obesity among Pakistani British children is consistent with NCMP surveillance^12^. Although excess weight became more apparent in mid-childhood within BMI-based surveillance systems, developmental origins research suggests that metabolic risk processes begin much earlier. Studies of early-life biomarkers, including leptin and insulin in cord blood, together with differences in infant feeding, diet, and physical activity, suggest that pathways to later obesity may originate in infancy and early childhood^24,25^. The mid-childhood divergence observed here may therefore reflect delayed manifestation of earlier biological and behavioural processes, although environmental and behavioural influences during mid-childhood may further amplify these susceptibilities. For example, previous BiB research shows declines in physical activity from around 7-8 years of age^26^.

#### Neurodevelopmental and common mental health conditions

Lower recorded rates of both CMHD and neurodevelopmental diagnoses among Pakistani British children are consistent with UK administrative and cohort analyses reporting lower diagnosed prevalence of autism spectrum disorder, ADHD, and some mental health conditions in minority ethnic groups compared with White British children^9,27^. However, the timing and likely mechanisms differ between these outcomes. CMHD emerged predominantly in adolescence, consistent with established developmental epidemiology of mood and anxiety disorders^28,29^. In contrast, neurodevelopmental conditions were identified earlier and showed a larger ethnic difference in recorded diagnoses. The neurodevelopmental composite was predominantly composed of socioemotional and behavioural diagnoses, consistent with autism spectrum disorder and ADHD, for which ethnic differences are known to be influenced by pathways to assessment, including referral practices, parental recognition, and access to specialist services^9,30,31^. Community-based surveys using standardised behavioural measurements have not consistently shown lower symptom prevalence, suggesting that differences in recorded diagnoses may also reflect variation in how symptoms are perceived, expressed, and responded to within families and communities^32,33^. Emerging evidence also suggests that familial support, community networks, and cultural interpretations of distress may influence symptom expression and help-seeking^6,34^. Lower recorded rates should therefore not be assumed to indicate lower underlying need.

#### Potential Mechanisms

Several mechanisms may underlie the observed ethnic differences. For atopic conditions, genetic susceptibility and early-life environmental exposures – including allergen burden and air pollution – have been implicated, and diagnostic practices may further shape recorded incidence. Within Bradford, air pollution, including traffic-related exposures, has been identified as an important contributor to childhood asthma risk^35^. Adiposity disparities likely reflect interactions between early growth patterns, diet, physical activity, and broader social determinants across childhood. Developmental origins research suggests that early-life programming and adiposity distribution may contribute to elevated cardiometabolic susceptibility in South Asian populations. Previous BiB research has shown that physical activity levels decline from around 7 – 8 years of age, with South Asian girls at particular risk^26^. Differences in neurodevelopmental and mental health diagnoses may reflect variation in cultural perceptions of developmental concerns, stigma, language barriers, help-seeking behaviour, and inequities in referral and assessment pathways^30,34,36^. Protective social and cultural factors – such as strong family support networks, community cohesion, and aspects of cultural identity – may also influence patterns of symptom expression and help-seeking and could contribute to lower recorded rates in some groups^37–39^. While our analyses were descriptive, the temporal patterning observed highlights critical windows for future causal investigation.

#### Strengths and Limitations

This study benefits from a large, ethnically diverse birth cohort with longitudinal linkage to primary care data, enabling follow-up from birth to adolescence. Use of consistent CTV3 diagnostic codes and a life-course framework allowed direct comparison of timing and trajectories across multiple conditions within the same population. However, routine primary care data capture diagnosed rather than symptomatic disease and may underestimate true prevalence, particularly for conditions subject to under-recognition or stigma. The classification of asthma within an atopic disease grouping may have introduced heterogeneity, as not all asthma is atopic in origin. The stepped appearance of cumulative incidence curves for overweight and obesity reflects the episodic BMI measurement rather than true discontinuities of risk. Neurodevelopmental conditions were identified using primary care diagnostic codes, which capture clinically recognised diagnoses but only one component of the broader identification pathway. Education-based classifications and validated behavioural instruments may identify additional cases. Future work linking healthcare, education, and cohort-derived measures would allow a more comprehensive assessment of developmental needs and potential disparities in diagnostic pathways. As this study was conducted within a single, socioeconomically deprived UK city with a high proportion of Pakistani-origin families, findings are most generalisable to similar urban populations and healthcare settings. Generalisability to other ethnic groups, less deprived populations, or non-UK healthcare systems may be limited.

#### Implications for Policy, Practice, and Research

The early emergence of atopic disparities suggests that prevention strategies should prioritise prenatal and early childhood periods in higher-risk communities. For obesity, early childhood is also a critical intervention window, although strategies must additionally address the amplification of risk during mid-childhood. Structural approaches targeting food environments, deprivation, and opportunities for physical activity are likely required alongside culturally tailored behavioural interventions. The contrasting pattern observed for neurodevelopmental and mental health diagnoses highlights the need to evaluate equity in access to assessment and care, while also considering whether protective social or cultural factors contribute to lower rates in some groups. Improved community engagement, culturally competent screening, and equitable referral pathways are essential. Future research should incorporate causal modelling and qualitative approaches to better understand biological, behavioural, structural, and healthcare-related mechanisms.

## Conclusions

This study shows that ethnic variation in childhood health differs by condition and timing. Pakistani British children experience a higher burden of early-onset atopic disease and later adiposity, whereas White British children have higher recorded rates of neurodevelopmental and mental health diagnoses. These differences highlight the importance of age-specific, culturally responsive approaches to prevention, service planning, and future research to understand underlying mechanisms.

## Data availability

Individual-level data from Born in Bradford study cannot be made publicly available due to ethical approvals and the need to protect participant confidentiality. Researchers may apply to access these data via the Born in Bradford Data Access process. Full details of the application process, eligibility criteria, and data access conditions are available at https://borninbradford.nhs.uk/our-data/how-to-access-data/. Applications are reviewed subject to appropriate scientific justification, governance approval, and completion of a data sharing agreement. Enquiries can be directed to the Born in Bradford research team.

## Supporting information

Supplementary Table S1: Asthma CTV3 codes

Supplementary Table S2: Atopic eczema CTV3 codes

Supplementary Table S3: Allergic rhinoconjunctivis CTV3 codes

Supplementary Table S4: Anxiety-related CTV3 codes

Supplementary Table S5: Depression-related CTV3

Supplementary Table S6: Neurodevelopmental disorder CTV3 codes

Supplementary Table S7: Numbers at risk

Supplementary Table S8: Neurodevelopmental subtypes

## Acknowledgements

Born in Bradford is only possible because of the enthusiasm and commitment of the children and parents in BiB. We are grateful to all the participants, health professionals, schools and researchers who have made Born in Bradford happen.

