## Supplementary Table S1: Asthma CTV3 codes for "Ethnic Differences in the Timing and Incidence of Childhood Health Conditions: Evidence from the Born in Bradford Cohort"

| **CTV3_CODE** | **Definition** |
| --- | --- |
| XaIR3 | Absent from work or school due to asthma |
| Xa9zf | Acute asthma |
| H333. | Acute exacerbation of asthma |
| Xafdj | Acute severe exacerbation of asthma |
| X101x | Allergic asthma |
| H33.. | Asthma |
| XaDvK | Asthma - currently active |
| XaDvL | Asthma - currently dormant |
| XaIIW | Asthma accident and emergency attendance since last visit |
| XaIeq | Asthma annual review |
| XE0YW | Asthma attack |
| XM0s2 | Asthma attack nos |
| XaINb | Asthma causes daytime symptoms 1 to 2 times per month |
| XaINc | Asthma causes daytime symptoms 1 to 2 times per week |
| XaINd | Asthma causes daytime symptoms most days |
| XaINZ | Asthma causes night symptoms 1 to 2 times per month |
| XaXZm | Asthma causes night time symptoms 1 to 2 times per week |
| XaXZp | Asthma causes symptoms most nights |
| 663N0 | Asthma causing night waking |
| XaJYe | Asthma clinical management plan |
| 1O2.. | Asthma confirmed |
| Y0024 | Asthma control |
| 8793. | Asthma control step 0 |
| 8794. | Asthma control step 1 |
| 8795. | Asthma control step 2 |
| 8796. | Asthma control step 3 |
| 8797. | Asthma control step 4 |
| 8798 | Asthma control step 5 |
| XaQHq | Asthma control test |
| XaIIZ | Asthma daytime symptoms |
| 663N. | Asthma disturbing sleep |
| 663N2 | Asthma disturbs sleep frequently |
| 663N1 | Asthma disturbs sleep weekly |
| XaIOV | Asthma finding |
| XaIer | Asthma follow-up |
| Y137e | Asthma guidance |
| 663P. | Asthma limiting activities |
| XaXZs | Asthma limits activities 1 to 2 times per month |
| XaXZu | Asthma limits activities 1 to 2 times per week |
| XaXZx | Asthma limits activities most days |
| XaINf | Asthma limits walking up hills or stairs |
| Xaam3 | Asthma management plan declined |
| 663x. | Asthma limits walking on the flat |
| 663U. | Asthma management plan given |
| XaIfK | Asthma medication review |
| 9OJ3. | Asthma monitor offer default |
| XM1Xb | Asthma monitoring |
| 9OJZ. | Asthma monitoring admin.nos |
| XaIu6 | Asthma monitoring by doctor |
| XaIu5 | Asthma monitoring by nurse |
| XaBU2 | Asthma monitoring call |
| 9OJ4. | Asthma monitoring call first letter |
| 9OJ5. | Asthma monitoring call second letter |
| 9OJ8. | Asthma monitoring call telephone invite |
| 9OJ6. | Asthma monitoring call third letter |
| 9OJ7. | Asthma monitoring call verbal invite |
| XE2Nb | Asthma monitoring check done |
| XaIRN | Asthma monitoring due |
| Xabj3 | Asthma monitoring invitation email |
| XacLz | Asthma monitoring sms text message 1st invitation |
| XacM0 | Asthma monitoring sms text message 2nd invitation |
| XacM1 | Asthma monitoring sms text message 3rd invitation |
| XaBU3 | Asthma monitoring status |
| Xabiu | Asthma monitorng invit sms (short message servce) txt messge |
| XaINa | Asthma never causes daytime symptoms |
| XaY2V | Asthma never causes night symptoms |
| 663O0 | Asthma never disturbs sleep |
| 663f. | Asthma never restricts exercise |
| XaIoE | Asthma night-time symptoms |
| XE0YX | Asthma nos |
| 663O. | Asthma not disturbing sleep |
| 663Q. | Asthma not limiting activities |
| 663W. | Asthma prophylactic medication used |
| Y0045 | Asthma prophylaxis drug id |
| 663e. | Asthma restricts exercise |
| XaX3n | Asthma review using roy colleg of physicians three questions |
| XaYb8 | Asthma self-management plan agreed |
| XaYZB | Asthma self-management plan review |
| 663e1 | Asthma severely restricts exercise |
| 663V. | Asthma severity |
| 663e0 | Asthma sometimes restricts exercise |
| XaIIX | Asthma treatment compliance satisfactory |
| XaIIY | Asthma treatment compliance unsatisfactory |
| XaIww | Asthma trigger |
| XaObi | Asthma trigger - airborne dust |
| XaLIr | Asthma trigger - animals |
| XaLJS | Asthma trigger - cold air |
| XaLJT | Asthma trigger - damp |
| Y01eb | Asthma trigger - dust mites |
| XaLJU | Asthma trigger - emotion |
| XaObj | Asthma trigger - exercise |
| YA598 | Asthma trigger - humidity |
| XaObk | Asthma trigger - pollen |
| XaLIm | Asthma trigger - respiratory infection |
| XaLIn | Asthma trigger - seasonal |
| XaObl | Asthma trigger - tobacco smoke |
| XaObm | Asthma trigger - warm air |
| YA599 | Asthma trigger - wind |
| Y0142 | Asthma trigger other |
| H33z. | Asthma unspecified |
| Xa0lZ | Asthmatic bronchitis |
| Ua1AX | Brittle asthma |
| XaIQ4 | Change in asthma management plan |
| X101t | Childhood asthma |
| X1027 | Colophony asthma |
| XaJ2A | Did not attend asthma clinic |
| 9OJ1. | Attends asthma monitoring |
| XaR8K | Did not attend asthma review |
| 8H2P. | Emergency admission, asthma |
| 663d. | Emergency asthma admission since last appointment |
| H335. | Chronic asthma with fixed airflow obstruction |
| Xa1hD | Exacerbation of asthma |
| XaJ50 | Excepted from asthma quality indicators: informed dissent |
| XaJ4z | Excepted from asthma quality indicators: patient unsuitable |
| XaJ4W | Exception reporting: asthma quality indicators |
| 173A. | Exercise-induced asthma |
| X1020 | Hay fever with asthma |
| 679J. | Health education - asthma |
| XaRFj | Health education - asthma self management |
| XaRFl | Health education - structured patient focused asthma discuss |
| Y139d | Indicators - asthma attack |
| H330. | Extrinsic (atopic) asthma |
| H330z | Extrinsic asthma NOS |
| H3301 | Extrinsic asthma with status asthmaticus |
| H3300 | Extrinsic asthma without status asthmaticus |
| Xafdz | Life threatening acute exacerbation of asthma |
| 663V1 | Mild asthma |
| 663V2 | Moderate asthma |
| XaLPE | Nocturnal asthma |
| XE0YT | Non-allergic asthma |
| XaINh | Number of asthma exacerbations in past year |
| 663V0 | Occasional asthma |
| H331. | Intrinsic asthma |
| H331z | Intrinsic asthma NOS |
| H3311 | Intrinsic asthma with status asthmaticus |
| H3310 | Intrinsic asthma without status asthmaticus |
| H33z2 | Late-onset asthma |
| XaRFi | Patient has a written asthma personal action plan |
| XaBAQ | Recent asthma management |
| H332. | Mixed asthma |
| 9OJ2. | Refuses asthma monitoring |
| XaNKw | Royal college of physicians asthma assessment |
| XaXa0 | Royal college physician asthma assessment 3 question score |
| 663V3 | Severe asthma |
| X102D | Status asthmaticus |
| H33z0 | Status asthmaticus NOS |
| XaIQE | Step down change in asthma management plan |
| XaIQD | Step up change in asthma management plan |
