## Supplementary Table S2: Atopic eczema CTV3 codes for "Ethnic Differences in the Timing and Incidence of Childhood Health Conditions: Evidence from the Born in Bradford Cohort"

| **CTV3_CODE** | **DESCRIPTION** |
| --- | --- |
| X505K | Eczema |
| XE1Av | Eczema NOS |
| M111. | Atopic dermatitis |
| M11.. | Atopic dermatitis and related conditions |
| M112. | Infantile eczema |
| M12z0 | Dermatitis NOS |
| XaINM | Exacerbation of eczema |
| XaY4o | Infected eczema |
| XE1C6 | Atopic eczema NOS |
| XaINK | Dry eczema |
| M113. | Flexural atopic dermatitis |
| Xa7lZ | Eczema of face |
| X505M | Constitutional eczema |
| Xa0p8 | Dermatitis of eyelid |
| XaY4Z | Infective eczematoid dermatitis |
| X505P | Discoid atopic dermatitis |
| X505R | Follicular atopic dermatitis |
| M11z. | Atopic dermatitis NOS |
| M114. | Allergic (intrinsic) eczema |
