## Supplementary Table S3: Allergic rhinoconjunctivis CTV3 codes for "Ethnic Differences in the Timing and Incidence of Childhood Health Conditions: Evidence from the Born in Bradford Cohort"

| **CTV3_CODE** | **DESCRIPTION** |
| --- | --- |
| Xa0lX | Seasonal allergic rhinitis |
| XE0Y5 | Allergic rhinitis |
| X00Zj | Seasonal allergic conjunctivitis |
| X00l9 | Hay fever unspecified allergen |
| H17z. | Allergic rhinitis NOS |
| X00Zk | Perennial allergic conjunctivitis |
| X1020 | Hay fever with asthma |
| XE16b | Other chronic allergic conjunctivitis |
| X00lA | Perennial allergic rhinitis |
| X00l8 | Hay fever other allergen |
| XE2QI | Allergic rhinitis due to pollens |
| H172. | Allergic rhinitis due to unspecified allergen |
| X00Zi | Atopic conjunctivitis |
| H170. | Pollinosis |
| XE0Y7 | Allergic rhinitis unspecified allergen |
| Hyu21 | [X]Other allergic rhinitis |
| F4C14 | Allergic conjunctivitis chronic |
| Hyu20 | [X]Other seasonal allergic rhinitis |
| X00lB | Perennial allergic rhinitis seasonal variation |
| XE0Y6 | Allergic rhinitis due to other allergens |
