## Supplementary Table S4: Anxiety-related CTV3 codes for "Ethnic Differences in the Timing and Incidence of Childhood Health Conditions: Evidence from the Born in Bradford Cohort"

| **CTV3_CODE** | **DESCRIPTION** |
| --- | --- |
| XE1aW | Anxiety states |
| .E31. | Pseudocyesis |
| E200. | Anxiety state |
| Eu41z | [X]Anxiety NOS |
| X00Sb | [X]Mild anxiety depression |
| .E4J6 | Anxiety with depression |
| E2003 | Anxiety with depression |
| Eu412 | [X]Mild anxiety depression |
| Xa7lj | Cancer phobia |
| E202B | Cancer phobia |
| E2028 | Claustrophobia |
| .E332 | Claustrophobia |
| E2004 | Chronic anxiety |
| .E311 | Chronic anxiety |
| E2020 | Phobia unspecified |
| E202C | Dental phobia |
| Eu400 | [X]Agoraphobia |
| X00SV | [X]Panic disorder+agoraphobia |
| .E331 | Agoraphobia |
| 1466 | H/O: anxiety state |
| E2001 | Panic disorder |
| XE1Y7 | Panic disorder |
| Ua1qS | Panic attack |
| .E310 | Panic attack |
| E200z | Anxiety state NOS |
| E2005 | Recurrent anxiety |
| .E312 | Recurrent anxiety |
| E2002 | Generalised anxiety disorder |
| Eu411 | [X]Anxiety reaction |
| Eu41. | [X]Other anxiety disorders |
| E2920 | Separation anxiety disorder |
| E2000 | Anxiety state unspecified |
| Eu410 | [X]Panic episodic paroxysm anx |
| X00T7 | [X]Anxious, avoidant personality |
| Eu606 | [X]Anxious, avoidant personality |
| Ub0qs | Anxiety management training |
| 8G94. | Anxiety management training |
| Eu40. | [X]Phobic anxiety disorders |
| Eu04. | [X]Acute psych-organic reaction |
| Eu402 | [X]Specific (isolated) phobias |
| X00SX | [X]Simple phobia |
| .E334 | School phobia |
| XE1YA | Phobic state |
| XM0Ak | School phobia |
| E202. | Phobic anxiety |
| X00SW | [X]Social neurosis |
| Eu401 | [X]Social neurosis |
| XaK2c | H/O: agoraphobia |
| 146G. | H/O: agoraphobia |
| .146G | H/O: agoraphobia |
| X00SY | [X]Needle phobia |
| .E335 | Needle phobia |
| Eu403 | [X]Needle phobia |
| E2022 | Agoraphobia - no panic attacks |
| 2258 | O/E - anxious |
| E2023 | Social phobia - eating in public |
| Eu515 | [X]Dream anxiety disorder |
| Eu930 | [X]Separation anxiety disorder childhood |
| E2025 | Social phobia - public washing |
| X00RP | [X]Organic anxiety disorder |
| Eu054 | [X]Organic anxiety disorder |
| E28z. | Flying phobia |
| X762H | Flying phobia |
| Eu41y | [X]Other specified anxiety disorder |
| XE1Zj | [X]Other specified anxiety disorder |
| Eu931 | [X]Phobic anxiety disorder childhood |
| Eu40y | [X]Other phobic anxiety disorder |
| E2026 | Acrophobia |
| XaL0q | Ref guide self-help for anxiety |
| 8HHp. | Ref guide self-help for anxiety |
| Eu932 | [X]Social anxiety disorder childhood |
| XE1aD | [X]Social anxiety disorder childhood |
| E2D0. | Anxiety/fear childhood/adolescence |
| E2024 | Social phobia - public speaking |
| Eu40z | [X]Phobic anxiety disorder unspecified |
| E2D0z | Anxiety/fear childhood/adolescence NOS |
| Eu413 | [X]Other mixed anxiety disorder |
| Eu93y | [X]Child overanxious disorder |
| 1B13. | Anxious |
| XE0rb | Anxious |
| XaboM | Ref guide self-help anxiety declined |
| 8IH53 | Ref guide self-help anxiety declined |
