## Supplementary Table S5: Depression-related CTV3 for "Ethnic Differences in the Timing and Incidence of Childhood Health Conditions: Evidence from the Born in Bradford Cohort"

| **CTV3_CODE** | **DESCRIPTION** |
| --- | --- |
| E2B.. | Depressive disorder NEC |
| Eu32z | [X]Depression NOS |
| E112. | Endogenous depression |
| X00Sb | [X]Mild anxiety depression |
| .E4J6 | Anxiety with depression |
| X00SQ | Agitated depression |
| .E221 | Mania/hypomania |
| E204. | Neurotic (reactive) depression |
| .E35. | Depression - postnatal |
| Eu32. | [X]Single episode depression |
| E290. | Brief depressive reaction |
| E11.. | Depressive psychoses |
| XE1aY | Postnatal depression |
| 1465 | H/O: depression |
| .62T1 | Puerperal depression |
| Eu530 | [X]Mild puerperal mental disorder |
| XE1Zb | [X]Depressive episode unspecified |
| XaB9J | Depression NOS |
| E2B0. | Postviral depression |
| Eu33. | [X]Recurrent depressive disorder |
| X761L | Seasonal affective disorder |
| E1137 | Recurrent depression |
| E2B1. | Chronic depression |
| X00SO | Depressed |
| XaCIu | Severe depression |
| Eu33z | [X]Recurrent depressive disorder unspecified |
| XE1Y0 | [X]Single episode depression |
| .E4JC | Recurrent depression |
| X00SS | Endogenous depression - first episode |
| Eu32y | [X]Other depressive episodes |
| X00SU | Masked depression |
| E113. | Endogenous depression recurrent |
| E112z | Single major depression NOS |
| Eu321 | [X]Moderate depressive episode |
| Eu322 | [X]Severe depressive episode no psychosis |
| Eu320 | [X]Mild depressive episode |
| Eu323 | [X]Severe depressive episode with psychosis |
| E291. | Prolonged depressive reaction |
| Eu333 | [X]Recurrent psychotic depression |
| XaLG0 | Depression resolved |
| X00S8 | [X]Post-schizophrenic depression |
| E11y2 | Atypical depressive disorder |
| Xa0Rd | Atypical depression |
| Eu331 | [X]Recurrent depression current moderate |
| Eu330 | [X]Recurrent depression current mild |
| XE1Zd | [X]Recurrent depression severe no psychosis |
| E1120 | Single major depression unspecified |
| E1130 | Recurrent major depression unspecified |
| E290z | Brief depressive reaction NOS |
| E02y3 | Drug-induced depressive state |
| Eu326 | [X]Major depression moderate severe |
| Eu325 | [X]Major depression mild |
| Eu327 | [X]Major depression severe without psychotic symptoms |
| XSGon | [X]Major depression severe with psychotic symptoms |
| XaY2C | Antenatal depression |
