## Supplementary Table S6: Neurodevelopmental disorder CTV3 codes for "Ethnic Differences in the Timing and Incidence of Childhood Health Conditions: Evidence from the Born in Bradford Cohort"

| **CTV3_CODE** | **DESCRIPTION** |
| --- | --- |
| YaxYA | Early childhood developmental disability |
| YaxYB | Early developmental impairment |
| Y01Ll | Infantile autism |
| Y01Lf | Autistic spectrum disorder |
| Y01Lh | Childhood autism |
| Y01Li | Kanner's syndrome |
| Y01Lq | Atypical autism |
| Y00Ly | Rett syndrome |
| Y01Ly | Asperger syndrome |
| Y01MW | Attention deficit hyperactivity disorder |
| Y01MV | Child attention deficit disorder |
| Y01MY | Attention deficit without hyperactivity |
| Y01Mq | Conduct disorder - unsocialised |
| Y01My | Conduct disorder - socialised |
| Y01N1 | Oppositional defiant disorder |
| YMB82 | Elective mutism |
| YMBzd | Selective mutism |
| Y01NB | Tic disorder unspecified |
| Y00K1 | Gilles de la Tourette syndrome |
| Y01LH | Disorder of speech and language development |
| Y01LO | Language development disorder |
| YMDry | Expressive language disorder |
| YMHib | Receptive language disorder |
| YMFPi | Global developmental delay |
| Y01LF | Mixed disorder of psychological development |
| Y01Ky | Mental retardation |
| Y01L0 | Mild mental retardation |
| Y01L5 | Moderate mental retardation |
| Y01L7 | Severe mental retardation |
| Y01L8 | Profound mental retardation |
