## Supplementary Table S7: Numbers at risk for "Ethnic Differences in the Timing and Incidence of Childhood Health Conditions: Evidence from the Born in Bradford Cohort"

| **Condition** | **Ethnicity** | **Age 0** | **Age 2** | **Age 4** | **Age 6** | **Age 8** | **Age 10** | **Age 12** | **Age 14** | **Age 16** |
| --- | --- | --- | --- | --- | --- | --- | --- | --- | --- | --- |
| **Allergic rhinoconjunctivitis** | Pakistani | 5878 | 5688 | 5422 | 5162 | 4967 | 4776 | 4660 | 3971 | 1522 |
|  | White British | 4818 | 4672 | 4420 | 4229 | 4069 | 3944 | 3826 | 3230 | 1237 |
| **Asthma** | Pakistani | 0 | 0 | 5462 | 5132 | 4929 | 4738 | 4621 | 3931 | 1525 |
|  | White British | 0 | 0 | 4397 | 4122 | 3921 | 3761 | 3611 | 3018 | 1158 |
| **Common mental health disorders** | Pakistani | 0 | 0 | 0 | 5617 | 5578 | 5511 | 5456 | 4668 | 1793 |
|  | White British | 0 | 0 | 0 | 4395 | 4289 | 4184 | 4055 | 3404 | 1292 |
| **Eczema** | Pakistani | 0 | 5686 | 5420 | 5160 | 4965 | 4774 | 4658 | 3969 | 1520 |
|  | White British | 0 | 4669 | 4417 | 4226 | 4066 | 3941 | 3823 | 3227 | 1234 |
| **Neurodevelopmental disorders** | Pakistani | 5878 | 5782 | 5664 | 5566 | 5514 | 5444 | 5394 | 4623 | 1801 |
|  | White British | 4818 | 4723 | 4519 | 4357 | 4218 | 4092 | 3931 | 3261 | 1248 |
| **Obesity** | Pakistani | 0 | 5054 | 4986 | 4741 | 4520 | 4210 | 3681 | 3127 | 1178 |
|  | White British | 0 | 3979 | 3899 | 3691 | 3499 | 3272 | 2893 | 2425 | 915 |
| **Overweight** | Pakistani | 0 | 4541 | 4452 | 4127 | 3870 | 3502 | 2860 | 2414 | 918 |
|  | White British | 0 | 3380 | 3265 | 2991 | 2812 | 2558 | 2161 | 1816 | 695 |
